# Hormone overdose and misuse in Chinese transgender and gender non-conforming population: A mixed-methods study protocol

**DOI:** 10.1101/2022.10.05.22280725

**Authors:** Ben-tuo Zeng, Hui-qing Pan, Li-ping Li, Tian-meng Lan, Zhen-yu Ye, Peng-fei Wang, Yang Liu

**Author notes:** Correspondence:* Yang Liu, School of Medicine, Xiamen University; 4221 Xiang’an Road (South), 361102 Xiamen, China. The authors contributed equally to this work (Ben-tuo Zeng, Hui-qing Pan). (B.-T.Z.); (L.-P.L.); (P.-F.W.). (H.-Q.P.). (T.-M.L.). (Z.-Y.Y.).

## Abstract

**Background:** There is no existing research on hormone overdose and misuse (HODM) in Chinese transgender and gender non-conforming (TGNC) population, and little is known in this field.

**Objectives:** We aim to determine the definition and criteria of HODM in Chinese TGNCs, address the rate of HODM in Chinese TGNC population, explore related factors and behavioral risks, identify the probable causes, and explore long-term effects.

**Methods:** We propose: (1) a mixed-method study comprising expert panel meetings and stakeholder engagement to identify HODM criteria, types and grades; (2) a cross-sectional study to quantify HODM prevalence, related factors and behavioral risks; (3) semi-structured interviews and focus groups to explore HODM motivations and reasons; and (4) a prospective cohort study to evaluate HODM long-term effects.

**Ethics:** The study protocol was approved by the Medical Ethics Committee of Xiamen University (XDYX202210K27).

**Dissemination:** Results will be published in international peer-reviewed journals, and a public-oriented version of the main findings will be prepared and disseminated through social media and online communities. The study will be completed before September 2023 except for the cohort study. Preliminary findings of the cohort study will be reported by March 2026.

## 1. Background & Rationale

Transgender and gender non-conforming (TGNC) individual is an umbrella term for those whose gender identity or gender expression differs from the assigned sex (or born sex). Gender affirming hormone therapy (GAHT), previously named transgender hormone replacement therapy (HRT), is one of the most preferrable approaches for TGNC individuals to change/modify their physiological sexual characteristics (PSC) and alleviate gender dysphoria (GD).

GAHT comprises several regimens of hormone use for different TGNC subgroups. Typical GAHT procedures include (a) estrogens and anti-androgens for transgender females (or male to female, MtF), (b) androgens for transgender males (or female to male, FtM), (c) gonadotropin releasing hormone analogues (GnRHa) for TGNC adolescents, and (d) modifiable hormone combinations^1^ for other gender non-conforming population to suppress/promote specific PSCs.

Standard GAHT under the supervision of clinicians and regular medical examinations is safe and effective for the change of PSCs and the remission of GD, even if the appropriate dose for a particular individual remains not clear enough^2^. Beyond normal doses of hormones, GAHT without supervision and medical examinations is harmful, with more side-effects, and can even reduce TGNC’s life expectancy. For example, long-term estrogen intake of high doses raises the cumulative dose of estrogen intake, hence increasing the risk of higher BI-RADS scores or even breast cancer of MtF^3^; and for FtM, polycystic ovary syndrome (PCOS) may be associated with high exogeneous androgen^4^. High hormone dosages also lead to hepatic dysfunction as the metabolic process takes place in the liver^2, 5^.

To date, multiple American and European associations have released various GAHT clinical guidelines with recommended indications, procedures, methods and doses of GAHT, i.e., WPATH 2012^6^, UCSF 2016^7^, and Endocrine Society 2017^8^. Within the guidelines, the recommended doses of hormones are clearly stated and the safety is guaranteed, despite without high-quality evidence. However, they are less accessible for Chinese TGNCs due to language barriers and medical knowledge levels.

Beyond the inaccessibility of online knowledge, it is also difficult for Chinese TGNCs to access TGNC healthcare resources. To our knowledge, there are extremely few TGNC-related endocrinologists and multidisciplinary treatment groups in China, where the only ones are all located in big cities, e.g., Beijing, Shanghai, and Guangzhou. Using the data from the US that there were 390 TGNCs per 100,000 US population in 2016^9^, it is estimated that there are 5.5 million TGNCs in China, indicating a serious disproportion between GAHT demands and healthcare resources. In this context, most Chinese TGNCs have to use hormones themselves without professional supervision, thus leading to health risks. Moreover, they have to access to hormones from irregular sellers without quality guarantees, such as personal drug agents.

Self-administration of hormones without essential medical knowledge bases can be very free but unsafe. From our previous interviews among online communities of TGNC individuals and the authors’ experiences as a transgender, the major of hormone administration-related knowledge of Chinese TGNC youths is from online forums and agents who illicitly sell hormones to TGNCs. This leads to unregulated GAHT regimens and use, where the most common condition is hormone overdose and misuse (HODM). Generally, HODM is a condition where TGNC individuals use sex hormones in excessive doses, or use hormone-like drugs which have been shown to be harmful with evidence, such as hexestrol. However, we still need to work on precise HODM definition.

There are few publications on HODM in GAHT process among TGNC individuals, where we have not retrieved any articles of this topic for Chinese TGNCs. The definition of HODM in Chinese TGNCs, related factors, causes, and long-term effects all need further investigation. It also remains unclear if the maximum safe dose and dose-response of hormones for Chinese TGNCs are the same as that for Caucasians/Latinos/Blacks, for whom the existing guidelines are mainly intended.

## 2. Aims & Research Questions

Our proposed study aims to investigate the definition of HODM in Chinese TGNCs, quantify the rate of HODM in Chinese TGNC population, related factors and risks, identify the probable causes of HODM, and follow up HODM individuals to explore long-term effects.

The research questions are:

*Q1* What is the definition and the criteria of HODM in Chinese TGNC population?

*Q2* What is the prevalence of HODM in Chinese TGNC population?

*Q3* What factors and risks are relevant to HODM?

*Q4* What are the causes?

*Q5* What are the long-term effects of HODM in Chinese TGNC population, compared to those who enroll in regular GAHT regimens?

## 3. Methods

### 3.1. Study Design

HODM is a complex sociomedical issue without existing clear criteria. Since there are no previous bases, it is hard to use quantitative methods only to evaluate the full picture. Therefore, we decide to employ a mixed-methods design to construct a full research framework on HODM.

The study can be divided into four stages to evaluate research questions (Table 1). Stage I will establish a precise definition and eligibility criteria of HODM through expert panel meeting and stakeholder engagement. Subsequently, a cross-sectional study will be conducted to evaluate the rate, subtypes, and related factors of HODM in Chinese TGNCs. In Stage III, semi-structured interviews and focus groups for TGNCs who are identified as HODM will be employed to investigate the causes, motivations and personal impact factors. This stage will be divided into two phases, Stage III-a before the cross-sectional study to provide a brief picture of HODM behaviors, and Stage III-b after the cross-sectional study to draw an overall pattern. Lastly, all participants of Stage II or III who would like to participate in our follow-up will be included in prospective cohorts to assess long-term effects.

**Table 1.** Study design.

| Stage | Qualitative / Quantitative | Study type | Research questions | Setting |
| --- | --- | --- | --- | --- |
| I | Qualitative | Expert panel meeting; Stakeholder engagement | Q1; Q3; Q4 | Online meeting |
| III-a | Qualitative | Semi-structured interview | Q3; Q4 | Online meeting |
| II | Quantitative | Cross-sectional study | Q2; Q3; Q4 | Online community and social media |
| III-b | Qualitative | Semi-structured interview; focus group | Q3; Q4 | Online meeting |
| IV | Quantitative | Prospective cohorts | Q3; Q4; Q5 | Online or via telephone |

### 3.2. Study Setting

We will conduct all stages of this study online due to the dispersed and small target population (Table 1). However, an offline meeting may be held in Stage I and Stage III if possible.

### 3.3. Study Procedure

#### 3.3.1. Stage I: Expert Panel Meeting and Stakeholder Engagement

Our purpose of Stage I is to identify HODM criteria, types and gradings, discuss the core scale to quantify HODM and related factors used in Stage II, and determine the primary and second outcomes used in Stage IV.

##### 3.3.1.1 Expert Panel Meeting

Since there are few researchers in the field of GAHT in China, it is not feasible to use a broad Delphi method to solicit expert opinions. Instead, we will organize panel meetings to discuss critical issues regarding HODM.

Researchers, endocrinologists, and clinicians specialized in the field of GAHT over the world will be invited to this panel meeting. All experts will have to report their conflicts of interest and sign informed consents before the meeting. The panel meeting will be held several times to adequately discuss all concerned topics (Table 2). Before and after the meetings, all panel members will receive several questionnaires on topics to discuss. A self-designed questionnaire on HODM according to the consensus will be disseminated to external experts after all panel meetings to solicit a broader range of opinions, and the final criteria will be decided after this survey.

**Table 2.** Expert Panel Meeting and Stakeholder Engagement Design

| Module | Participants | Requirements | Plan | Topics to Discuss |
| --- | --- | --- | --- | --- |
| Expert Panel Meeting | Researcher; Endocrinologist; Clinician; Statistician; Methodologist | Conflict of Interest Form; Written informed consent | Three meetings plus e-mail communication and several questionnaires. | a. Definition of HODM in each TGNC subtype<br>b. The appropriateness of existing recommended dosages for Chinese population |
|  |  |  | All in English. | c. Criteria for HODM, including dosages, duration, frequency, and hormone types<br>d. Types and grades of HODM<br>e. Related factors and probable causes of HODM<br>f. Essential items for the main scale<br>g. Outcomes for the prospective cohorts |
| Stakeholder Engagement | TGNC;<br>Parent of TGNC;<br>Psychiatrist;<br>Psychological counsellor;<br>Plastic surgeon;<br>Primary care clinician;<br>Other related professionals | Conflict of Interest Form;<br>Written informed consent | Three to four meetings plus a questionnaire.<br>In English and Chinese. | a. Definition of HODM in each TGNC subtype<br>b. Is this term offensive?<br>c. Routine hormone dosages<br>d. Criteria, types and grades for HODM (as a complement to expert panel meeting)<br>e. HODM common situations<br>f. Related factors and probable causes of HODM<br>g. Motivation and triggers of HODM |

##### 3.3.1.2 Stakeholder Engagement

Stakeholder engagement will be held after expert panel meetings to collect more information on possible relevant factors and causes, and complement the definitions and criteria identified by experts.

Existing stakeholder engagement guidance in healthcare fields are mainly for clinical guidelines or systematic reviews, but not mixed methods studies. Therefore, we adjust methods from a six-step stakeholder engagement framework^10^ from Cochrane for our study (Figure 1).

**Figure 1.**
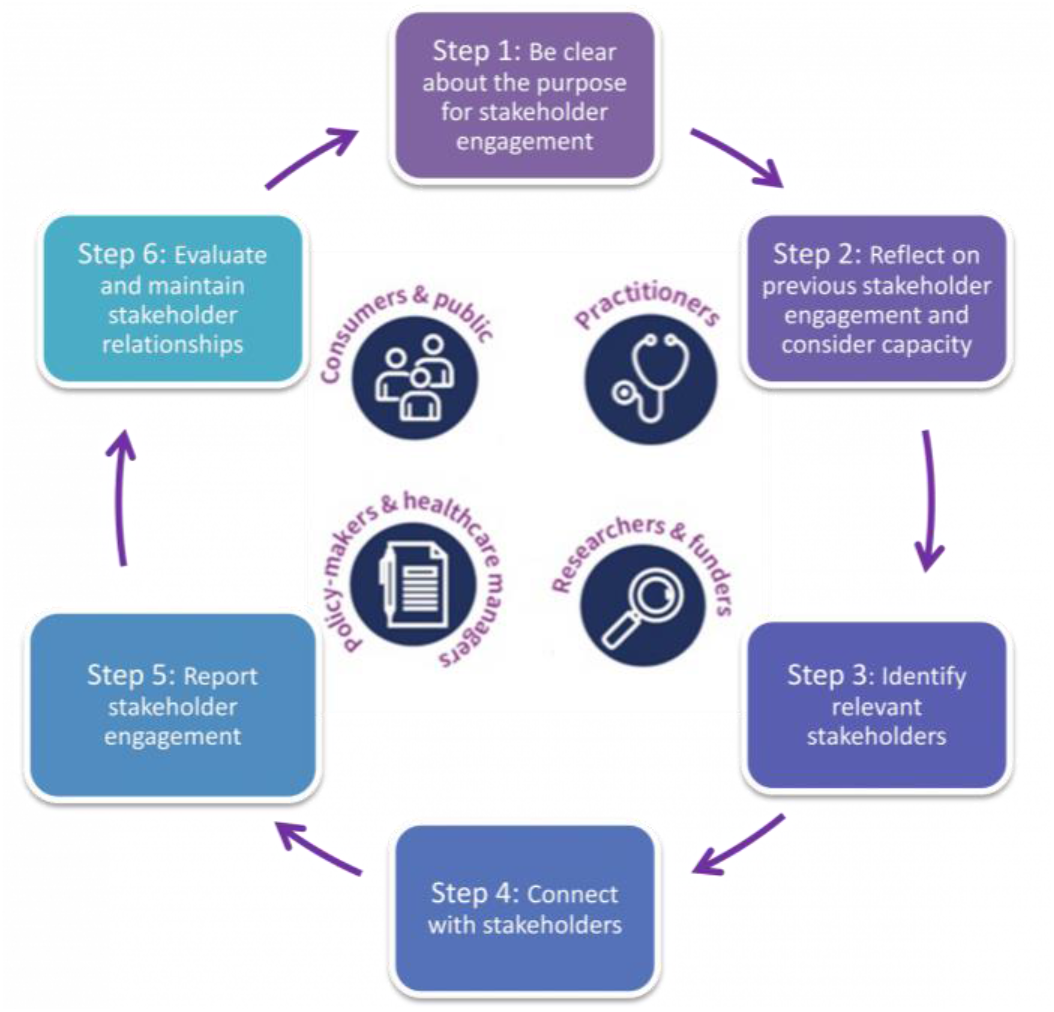
Stakeholder engagement framework^10^. This is not an original image. The original image is available at: https://training.cochrane.org/online-learning/knowledge-translation/meaningful-partnerships/engaging-stakeholders

We will invite five to fifteen Chinese TGNC individuals with GAHT experiences, including MtF, FtM and gender non-conforming/queer/non-binary individuals to participate in. We will also invite external experts in the field of TGNC from within and outside China, comprising primary care clinicians, psychiatrists, psychological counsellors, and plastic surgeons. These professionals are not specialized with GAHT but associated with TGNCs. Parents of TGNC individuals may also participate.

The stakeholder engagement meeting will be held three to four times and in Chinese and English separately to fit each participant’s timeline and language. Before the meetings, each stakeholder should report conflict of interest and sign an informed consent. We will make an outline prior to the meeting, comprising following elements:

a. an introduction to the study and the research group;
b. how we use the contents of the meeting;
c. an overview of previous meetings;
d. an introduction to current recommended hormone dosages;
e. questions to discuss (see Table 2);
f. time for stakeholders to identify other relevant factors, experience, or considerations;
g. a brief summary.

#### 3.3.2. Stage II: Cross-sectional Study

##### 3.3.2.1 Questionnaire design

We aim to quantify the rate of HODM, types and gradings of HODM, related behavioral features, and HODM behavioral risks through Stage II. We will design a questionnaire to evaluate HODM in Chinese TGNC population and related factors since there are not relevant publications.

The questionnaire will need 10 to 15 minutes to complete and comprise four sections: (1) demographics; (2) medical history, GAHT-related physical changes, and adverse effects; (3) a scale to evaluate HODM and related factors; and (4) other relevant items identified through Stage I. At the last of questionnaire, participants will be asked if they would like to participate in following stages (interviews, focus groups and cohorts).

The main scale in section 3 will contain three parts: (1) GAHT medication details and related information; (2) a scale to evaluate the severity of HODM; and (3) a score to measure HODM behavior risks and the possibility to change based on Theoretical Domains Framework^11^ version 2^12, 13^ (TDFv2). There are fourteen domains and 84 theoretical constructs within TDFv2, to clearly identify influences on behaviors and behavioral changes (Table 3).

**Table 3.**
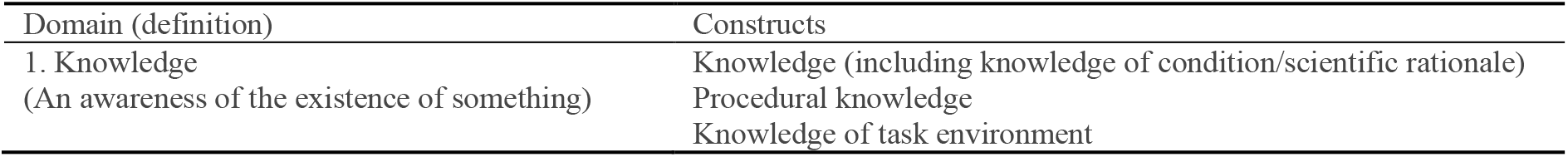
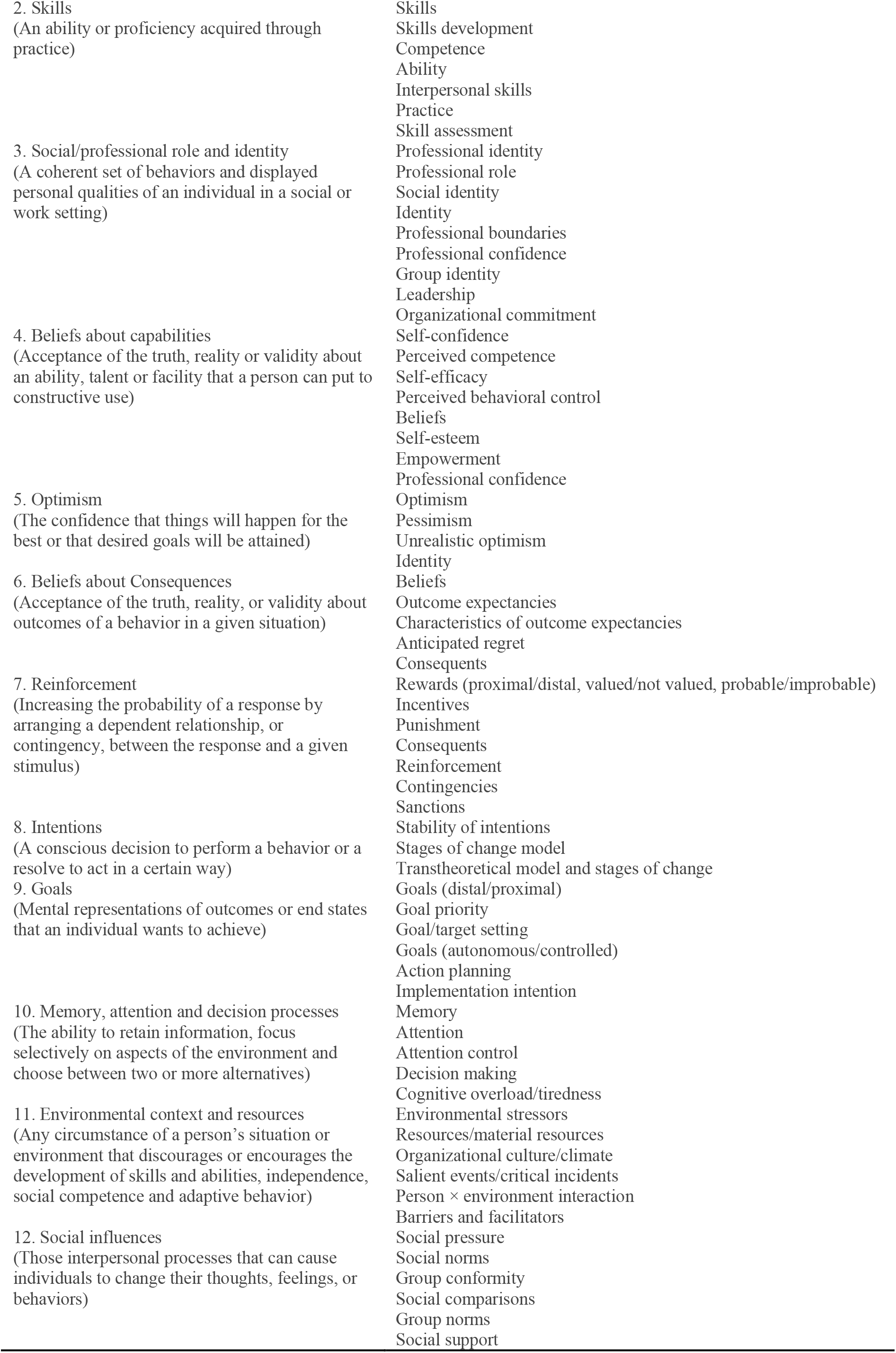
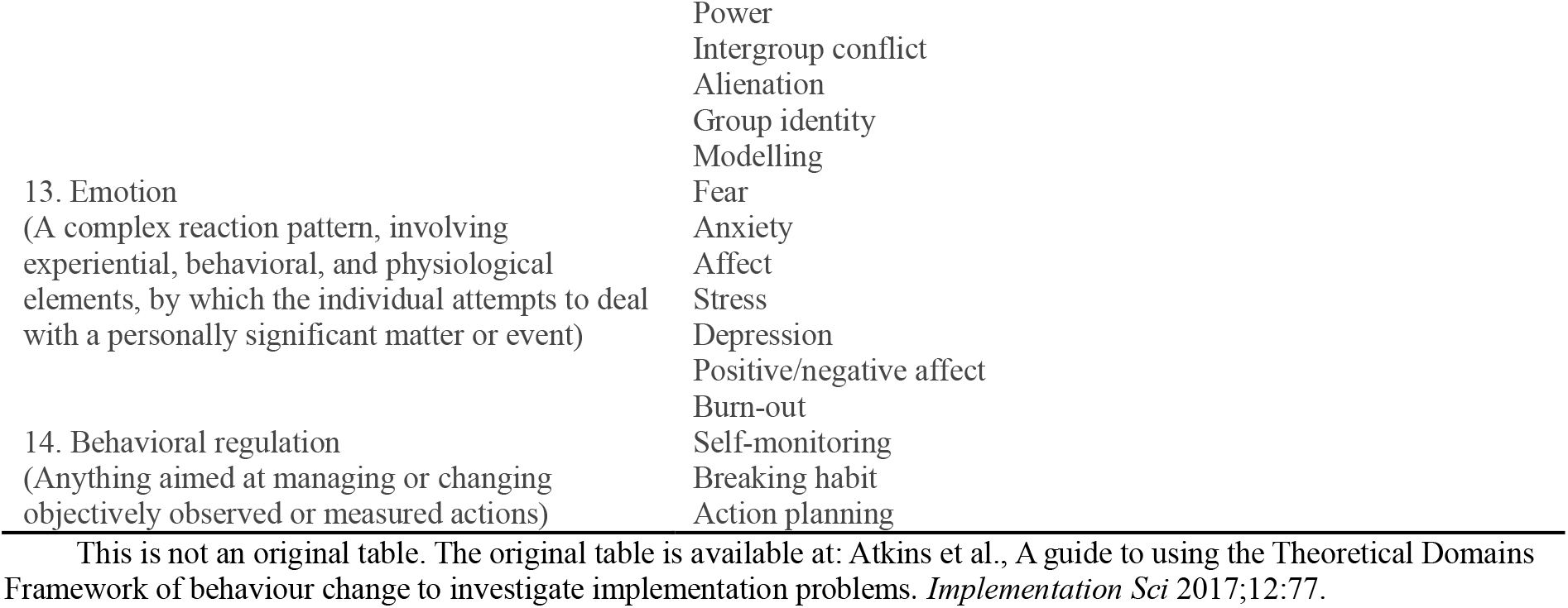
Theoretical Domains Framework version 2^12^

| Domain (definition) | Constructs |
| --- | --- |
| 1. Knowledge<br>(An awareness of the existence of something) | Knowledge (including knowledge of condition/scientific rationale)<br>Procedural knowledge<br>Knowledge of task environment |
| 2. Skills<br>(An ability or proficiency acquired through practice) | Skills<br>Skills development<br>Competence<br>Ability<br>Interpersonal skills<br>Practice<br>Skill assessment |
| 3. Social/professional role and identity<br>(A coherent set of behaviors and displayed personal qualities of an individual in a social or work setting) | Professional identity<br>Professional role<br>Social identity<br>Identity<br>Professional boundaries<br>Professional confidence<br>Group identity<br>Leadership<br>Organizational commitment |
| 4. Beliefs about capabilities<br>(Acceptance of the truth, reality or validity about an ability, talent or facility that a person can put to constructive use) | Self-confidence<br>Perceived competence<br>Self-efficacy<br>Perceived behavioral control<br>Beliefs<br>Self-esteem<br>Empowerment<br>Professional confidence |
| 5. Optimism<br>(The confidence that things will happen for the best or that desired goals will be attained) | Optimism<br>Pessimism<br>Unrealistic optimism<br>Identity |
| 6. Beliefs about Consequences<br>(Acceptance of the truth, reality, or validity about outcomes of a behavior in a given situation) | Beliefs<br>Outcome expectancies<br>Characteristics of outcome expectancies<br>Anticipated regret<br>Consequents |
| 7. Reinforcement<br>(Increasing the probability of a response by arranging a dependent relationship, or contingency, between the response and a given stimulus) | Rewards (proximal/distal, valued/not valued, probable/improbable)<br>Incentives<br>Punishment<br>Consequents<br>Reinforcement<br>Contingencies<br>Sanctions |
| 8. Intentions<br>(A conscious decision to perform a behavior or a resolve to act in a certain way) | Stability of intentions<br>Stages of change model<br>Transtheoretical model and stages of change |
| 9. Goals<br>(Mental representations of outcomes or end states that an individual wants to achieve) | Goals (distal/proximal)<br>Goal priority<br>Goal/target setting<br>Goals (autonomous/controlled)<br>Action planning<br>Implementation intention |
| 10. Memory, attention and decision processes<br>(The ability to retain information, focus selectively on aspects of the environment and choose between two or more alternatives) | Memory<br>Attention<br>Attention control<br>Decision making<br>Cognitive overload/tiredness |
| 11. Environmental context and resources<br>(Any circumstance of a person's situation or environment that discourages or encourages the development of skills and abilities, independence, social competence and adaptive behavior) | Environmental stressors<br>Resources/material resources<br>Organizational culture/climate<br>Salient events/critical incidents<br>Person × environment interaction<br>Barriers and facilitators |
| 12. Social influences<br>(Those interpersonal processes that can cause individuals to change their thoughts, feelings, or behaviors) | Social pressure<br>Social norms<br>Group conformity<br>Social comparisons<br>Group norms<br>Social support |
|  | Power |
|  | Intergroup conflict |
|  | Alienation |
|  | Group identity |
|  | Modelling |
| 13. Emotion<br>(A complex reaction pattern, involving experiential, behavioral, and physiological elements, by which the individual attempts to deal with a personally significant matter or event) | Fear |
|  | Anxiety |
|  | Affect |
|  | Stress |
|  | Depression |
|  | Positive/negative affect |
|  | Burn-out |
| 14. Behavioral regulation<br>(Anything aimed at managing or changing objectively observed or measured actions) | Self-monitoring |
|  | Breaking habit |
|  | Action planning |

This is not an original table. The original table is available at: Atkins et al., A guide to using the Theoretical Domains Framework of behaviour change to investigate implementation problems. *Implementation Sci* 2017;12:77.

We will develop the main scale after Stage I, and test for reliability and validity through a preliminary survey. The proposed items are listed in Table 4.

**Table 4.** Proposed items in the questionnaire.

| Variables | Type | Operationalization/Data range |
| --- | --- | --- |
| *Section 1: Demographics* |  |  |
| Age | D | 12 to 80 |
| Region | N | Province – Region (SW/SE/NW/NE China) |
| Assigned sex | N | Female/Male/Intersex/Other |
| Gender identity/expression | N | Female/Male/Non-binary/Cross-dressing/Not sure/Other |
| Sex orientation | N | Homosexual/Heterosexual/Pansexual/Bisexual/Asexual/Other |
| Ethnicity | N | Hans/Other ethnic minorities |
| Education level | O | Primary/Secondary/Upper secondary/Bachelor's/Master/Doctor |
| Major (if applicable) | S | (Flexible) |
| Employment | N | Student without part-time job/Student with part-time job/Part-time job/Full-time job/Unemployed/Self-employed/Seeking opportunities/Retired |
| Occupation (if applicable) | S | (Flexible) |
| Personal income | O | None/0 – 2,000/2,000 – 5,000/5,000 – 8,000/8,000 – 12,000/12,000–20,000/>20,000 CNY per month |
| Marital status | N | Not married/Married/Divorced/Widowed |
| Other spousal relationships | N | One spouse/two or more spouses/No spouse |
| Living status | N | Renter, self/Renter, co-renting/Homeowner/Live with family/Student dormitory/Company dormitory/Other |
| Religion | N | None/Buddhism/Taoism/Catholicism/Protestantism/Islam/Other |
| *Section 2: Medical history, GAHT-related physical changes, and adverse effects* |  |  |
| Height | C | in centimeters |
| Weight | C | in kilograms |
| Previous physical disorders | N | Yes/No |
| -Types | N | ICD-11 classification list<sup>14</sup> |
| Current physical disorders (within 6 months) | N | Yes/No |
| -Types | N | ICD-11 classification list |
| Previous mental disorders | N | Yes/No |
| -Types | N | ICD-11 classification list |
| -Names | N | ICD-11 classification list |
| Current mental disorders (within 1 year) | N | Yes/No |
| -Types | N | ICD-11 classification list |
| -Names | N | ICD-11 classification list |
| Regular medical examination | N | Yes/No |
| -Frequency | C | ... (with units) |
| Gender Affirming Surgery | N | Yes/No/In progress |
| Physical changes and adverse |  |  |
| effects |  |  |
| -Fatigue | O | 5-point scale |
| -Nausea | O | 5-point scale |
| -Sex arousal | O | 5-point scale |
| -... | O | Other items to be addressed |
| *Section 3: Core Scale to evaluate HODM\** |  |  |
| 3.1 GAHT medication in the last year |  |  |
| -Types | N | Estrogen only/Estrogen and progesterone complex/Anti-androgen/Androgen/GnRHa/Progesterone/Anti-estrogen (estrogen receptor blocker/Other |
| -Name of agents | S | (Flexible) |
| -Routes of Administration | N | p.o./i.m./s.c./transdermal/other |
| -Frequency | N | b.i.d./q.d./q.wk./q.2wk./q.m.t./per ... month/artificial cycle |
| -Routine dose | C | ... (with units) |
| -Minimal dose (once) | C | ... (with units) |
| -Maximal dose (once) | C | ... (with units) |
| -Frequency of maximal dose | C | ... (within one year) |
| 3.2 Serum sex hormones (if applicable, the latest data) |  |  |
| -Estradiol | C | ... (with units) |
| -Estriol/Estrone | C | ... (with units) |
| -Testosterone | C | ... (with units) |
| -Progesterone | C | ... (with units) |
| -Prolactin | C | ... (with units) |
| -Gonadotropins | C | ... (with units) |
| 3.3 Other Information: |  |  |
| Duration of GAHT | C | ... (years, months) |
| Age when first GAHT | D | 12 to 80 |
| Interruptions | N | Yes/No |
| -Duration | C | ... (years, months) |
| Medication Change | N | Yes/No |
| -Details | S | ... |

D, discrete; N, nominal; O, ordinal; S, character strings; C, continuous
\*: Only GAHT-related basic items are listed here. The full version of the scale will be developed after Stage I.

**Table 4.** Themes for semi-structured interviews and focus groups

| Phase | Target interviewer | Themes |
| --- | --- | --- |
| Stage III-a | All TGNCs with GAHT experience (semi-structured interview) | a. GAHT history, experiences, and motivations<br>b. GAHT-related knowledge sources<br>c. HODM-related knowledge and experiences<br>d. Influence of GAHT and HODM |
| Stage III-b | Non-HODM TGNCs (semi-structured interview) | a. Current GAHT regimens<br>b. Interpretation on HODM<br>c. Experiences with HODM ideas<br>d. Potential risks on HODM<br>e. Why do some TGNCs have HODM? |
|  | HODM TGNCs (semi-structured interview) | a. Do you think you have had HODM?<br>b. Motivations for HODM<br>c. Incentives and triggers for HODM<br>d. Interpretation on HODM |
|  | Severe HODM focus group | a. Frequency and severity of HODM<br>b. Experience and Feelings about HODM<br>c. External and internal factors prompting HODM<br>d. Other causes for HODM |
|  | High behavioral risk focus group | a. The role of personal experience in HODM<br>b. The role of external environments in HODM<br>c. The role of mental disorders and gender dysphoria in HODM<br>d. How can we act to reduce the risk? |

Within section 4, according our previous interviews, we will use PHQ-9^15^, GAD-7^16^, and **Utrecht Gender Dysphoria Scale – Gender Spectrum (UGDS-GS)**^**17**^ to assess depression, anxiety, and gender dysphoria, respectively. Moreover, Body Esteem Scale (BES)^18^ and **Transgender Congruence Scale (TCS)**^**19**^ will be employed to assess TGNC’s body image and transgender congruence. Other items will be considered if the experts and stakeholders agree they are critical, while the mentioned scales may be deleted if the experts consider them as misappropriate. If validated Chinese versions of included scales are available, we will request the Chinese versions and use them; otherwise, we will translate scales into Chinese and validate them.

##### 3.3.2.2. Eligibility Criteria for Participants

The inclusion criteria for this cross-sectional study will be:

a. individuals with Chinese nationality (excluding Hong Kong SAR, Macau SAR, and Taiwan because of different social backgrounds);
b. aged over 12, since there will be difficulty understanding the questions for those under 12 years old, and the youngest adolescents who began GnRHa treatment was typically 11 to 12 years old from previous trials^20, 21^;
c. identify themselves as TGNCs of any subtypes.
d. have experienced at least one month of GAHT of any regimens and frequency in the last one year, or is taking hormones during the period of the study.
e. Hormones taken include androgen, estrogen, progesterone, anti-androgen, and anti-estrogen of any chemical structures, trade names, or regimens.

The exclusion criteria will be:

a. individuals who undertake GAHT in the absence of self-consciousness due to mental disorders, or under external coercion/abuse;
b. refuse to sign or unable to understand the informed consent;
c. other considerations to be identified via Stage I.

##### 3.3.2.3. Pre-test, Sample Size, and Sampling

We will conduct a small-scale pre-test online to validate scales and calculate information for sample size calculation. Considering the difficulty to collect sample, the proposed size for pre-test will be approximately 50 as previously estimated and according to current trials in the field of GAHT^8^.

Because of the dispersed and rare population, we will use exponential non-discriminative snowball sampling strategy^22, 23^ to cover a wide range of TGNCs from across China and obtain sufficient participants. We will disseminate the questionnaire among online TGNC communities, social medias, under the help TGNC-related non-government organizations (NGO). Each participant will be asked to share the questionnaire link to other TGNCs.

In 2017, a nation-wide TGNC survey by one of the largest LGBT NGO in China garnered a sample of 2,060 with a response rate of 36.30%, where around one third of the participants had experienced GAHT^24^. Given the data, it is estimated that there are 1.71 million GAHT-experienced TGNCs in China. Assuming a prevalence of 0.50 for HODM, a 95% confidence interval width of 0.10, and a response rate of 0.80, it is estimated that the sample size should be no less than 482 using PASS 15.0 (NCSS LLC., UT, USA).

#### 3.3.3. Stage III: Semi-structured Interview and Focus Group

##### 3.3.3.1 Participants Selection

Interviewees for Stage III-a will be selected using the same eligibility criteria for Stage II, and all participants of Stage III-b will be recruited from within the sample of Stage II. Only those who answer “Yes” for their intention to participate in Stage III-b will be considered. However, if there are few eligible interviewees, we may recruit participants through other processes.

Through Stage I, HODM will be graded as from mild to severe, and HODM-related behavioral factors will be graded as from low-risk to high-risk accordingly. Different types of HODM will also be identified. We will select participants according to their types and grades of HODM during Stage III-b. HODM and non-HODM individuals identified by the questionnaire will be included in semi-structured interview, while there will be two focus groups for HODM participants with severe HODM and high-risk behavioral factors. The number of interviewees depends on the sample size of Stage II. If possible, we estimate that there will be 4 to 10 persons per focus group, and we will recruit up to 30 non-HODM and HODM participants for semi-structured interview. As for Stage III-a, only semi-structured interviews will be implemented. We will not distinguish between HODM and non-HODM individuals since the survey has not been conducted.

##### 3.3.3.2 Themes

Themes to discuss in this stage are displayed in Table 4.

#### 3.3.4. Stage IV: Prospective Cohorts

We aim to evaluate long-term effects of HODM in a prospective cohort study over a 2-to 4-year period.

##### 3.3.4.1. Recruitment, Eligibility Criteria, and Sample Size

The eligibility criteria for TGNCs to participate in this stage is the same as that in Stage II. All TGNCs who participate in Stage II or Stage III and agree to be followed up will be eligible for this stage, while if needed we will also recruit more TGNCs from outside previous participants via social media. It is difficult to determine appropriate sample size before Stage II.

##### 3.3.4.2. Allocation

We will divide participants into two cohorts, those with HODM (cohort 1) and without HODM (cohort 2).

##### 3.3.4.3. Endpoints and Measurements

Our proposed endpoints will be: (1) adverse events (Grade 2 or higher in CTCAE^25^); (2) GAHT attrition due to adverse events; (3) gender dysphoria, depression, and anxiety; (4) quality of life; (5) Body image and body-gender congruence; (6) Tanner stages and changes; (7) Change of gender identity; and (8) Other endpoints to be identified through Stage I. The priority of endpoints needs further discussion in expert panel meetings.

Measurements of these endpoints will be: (respectively) (1) self-reported and classified using CTCAE v5.0; (2) self-reported; (3) PHQ-9^15^, GAD-7^16^, and UGDS-GS^17^; (4) 36-item short form health survey (SF-36)^26^; (5) BES^18^ and TCS^19^; (6) visual analogue scale (VAS); and (7) self-reported.

We will measure all the outcomes above at 0, 1, 3, 6, 12, 18, 24, 30, 36, 42, 48 months after the baseline assessment. Additionally, participants can report their adverse events, GAHT attrition, and change of gender identity at any time during the follow-up.

### 3.4. Data Analysis

#### 3.4.1. Qualitative Data

In Stage I, we will use SurveyPlanet (SurveyPlanet LLC., DC, USA) for conflict-of-interest forms and other questionnaires. All meetings and interviews in Stage I and III will be video-taped and transcribed. For Stage I, we will re-play the records to ensure that all related information has been extracted. For Stage III, Colaizzi’s seven-step method^27^ will be used. Transcripts and observation narratives will be imported into NVivo 20 (QSL International, MA, USA) for coding. Two researchers will independently identify themes according to the outlines and topics used during interviews.

#### 3.4.2. Cross-sectional Study

We will use Tencent Survey (Tencent Cloud Computing, Beijing, China) platform for data collection. Data will be imported to MySQL 8.0 (Oracle Corporation, TX, USA) for management. We will use R 4.2.1 (The R Foundation, Vienna, Austria) for statistical analysis.

For psychometrics of the scales included in our questionnaire, we will test for Internal consistency reliability and report Cronbach’s α coefficient, where 0.70 is the threshold. As for validity, Kaiser-Meyer-Olkin (KMO), Bartlett test, and exploratory factor analysis will be applied. The threshold for KMO test will be 0.60.

We will present descriptive data characteristics firstly, and test for normality of continuous variables. Subsequently, we will estimate the rate of HODM among Chinese TGNC population, and test for differences between HODM participants and non-HODM participants using independent *t*-test, *ANOVA, χ*^2^ test, or non-parametric test regarding variable types. Correlation analysis will be used among HODM, behavioral score, and other variables to explore correlations. Finally, univariate and multivariate logistic regression will be applied. Before regression, we will use directed acyclic graphs (DAGs) to exclude potential confounders. Normally distributed data will be presented as mean and standard deviation, while skewed data will be displayed as median and interquartile range. 95% confidence interval will be used for all inferential data.

#### 3.4.3. Prospective Cohorts

Outcomes will be present as risk ratio (RR), mean difference (MD), or hazard ratio (HR), and 95% confidence intervals, regarding variable types. We will adjust outcomes through univariate and multivariate analysis. Confounders will be identified using DAGs and through univariate analysis.

## 4. Ethical Considerations & Dissemination

The study protocol was approved by the Medical Ethics Committee of Xiamen University. All procedures of this study will follow the Declaration of Helsinki^28^. Individuals who participate in any stages of the study will have to provide a written informed consent if available. For online survey, participants can only access the questionnaire after clicking the “I have read and agree” button below all informed items.

Regarding data confidentiality, we use strong password for all related online platforms, and the username and password will only be shared within authors. Any sensitive information including address, IP address, legal names, and other identifiers will not be collected, except for contact details for those who agree to participate in following stages. In case sensitive information is collected by the survey tools automatically, we will delete them from downloaded data. After the data collection phase, all data will be deleted from online platforms. Data files will be stored on the local disk only, and will only be shared among authors using email if necessary.

We will publish the results in international peer-reviewed journals, and will present findings at academic conferences if possible. We will also prepare articles and reports of main findings in Chinese and disseminate them through social media and online communities. The study will start in October 2022, and Stage 1-3 of this study will be completed by September 2023. Preliminary results of the final stage (prospective cohorts) will be reported by March 2026.

## Data Availability

This is a protocol and does not produce any data.

## Supplemental Material

Informed Consents (in Chinese) and Ethical Approval (in Chinese).

